# Oncologic Immunomodulatory Agents in Patients with Cancer and COVID-19

**DOI:** 10.1101/2020.08.11.20145458

**Authors:** Justin Jee, Aaron J. Stonestrom, Sean Devlin, Teresa Nguyentran, Beatriz Wills, Varun Narendra, Michael B. Foote, Melissa Lumish, Santosha Vardhana, Stephen M. Pastores, Neha Korde, Dhwani Patel, Steven Horwitz, Michael Scordo, Anthony Daniyan

**Author notes:** Co-senior authors (Michael Scordo, Anthony Daniyan). Corresponding authors: 1275 York Avenue, New York, NY 10065, 1275 York Avenue, New York, NY 10065.

## Abstract

**Background:** Corticosteroids, anti-CD20 agents, immunotherapies, and cytotoxic chemotherapy are commonly used in the treatment of patients with cancer. How these agents impact patients with cancer who are infected with SARS-CoV-2 remains unclear.

**Methods:** We retrospectively investigated associations between SARS-CoV-2-associated respiratory failure or death with receipt of the aforementioned medications and with pre-COVID-19 neutropenia. The study included all cancer patients diagnosed with SARS-CoV-2 at Memorial Sloan Kettering Cancer Center until June 2, 2020 (N=820). We controlled for cancer-related characteristics known to predispose to worse COVID-19. To address that more acutely ill patients receive therapeutic corticosteroids, we examined patient subsets based on different levels of respiratory support: ≤2 L/min supplemental oxygen, >2L/min supplemental oxygen, and advanced respiratory support prior to death.

**Results:** Corticosteroid administration was associated with worse outcomes in the pre-2L supplemental oxygen cohort; no statistically significant difference was observed in the >2L/min supplemental oxygen and post-critical cohorts. Interleukin-6 (IL-6) and C-reactive protein (CRP) levels were lower, and ferritin levels were higher, after corticosteroid administration. In patients with metastatic thoracic cancer, 9 of 25 (36%) and 10 of 31 (32%) had respiratory failure or death among those who did and did not receive immunotherapy, respectively. Seven of 23 (30%) and 52 of 187 (28%) patients with hematologic cancer had respiratory failure or death among those who did and did not receive anti-CD20 therapy, respectively. Chemotherapy itself was not associated with worse outcomes, but pre-COVID-19 neutropenia was associated with worse COVID-19 course. Relative prevalence of chemotherapy-associated neutropenia in previous studies may account for different conclusions regarding the risks of chemotherapy in patients with COVID-19. In the absence of prospective studies and evidence-based guidelines, our data may aid providers looking to assess the risks and benefits of these agents in caring for cancer patients in the COVID-19 era.

## Introduction

The practical management of Coronavirus Disease 2019 (COVID-19), caused by severe acute respiratory syndrome coronavirus 2 (SARS-CoV-2), and management of cancer in SARS-CoV-2 endemic regions remains a major challenge for oncologists. Severe COVID-19 is characterized by a profound and uncontrolled pro-inflammatory cytokine response that can lead to lung injury and death [1]. However, the risk profile of immunomodulatory agents commonly used in standard cancer management is not established in patients with COVID-19. Recent studies [2-7] have identified cancer patient characteristics, such as thoracic and hematologic malignancy, associated with worse outcomes, but there is mixed data regarding the outcomes of cancer patients with COVID-19 treated with immunomodulatory agents.

For instance, the World Health Organization (WHO) and Infectious Disease Society of America (IDSA) initially recommended against the routine use of corticosteroids in patients with COVID- 19, in part due to concerns that they may decrease viral clearance, and only recently did the IDSA suggest corticosteroid use in hospitalized patients with severe COVID-19 based on new evidence of potential benefit in noncancer patients [9-12]. Corticosteroids are common components of cancer treatment and supportive care. However, their impact on cancer patients with COVID-19 is not well studied. A recent retrospective study found a possible trend toward worse outcomes associated with corticosteroid use in cancer patients, although no analysis was performed to correct for possible selection bias in which sicker patients received those medications [11].

Recent evaluations of the impact of cytotoxic chemotherapy on COVID-19 outcomes have found differing results [4,5,7,8]. Although in a 309-patient study, we found no evidence of worse outcomes in patients who had received recent cytotoxic chemotherapy in general, we did find that pre-COVID-19 neutropenia was associated with worse COVID-19 outcomes [7]. Further analysis regarding the interactions between neutropenia and chemotherapy at the time were precluded by sample size. Observational studies have also reported variable results regarding the impact of immunotherapy (i.e. PD-1 and CTLA-4 blockade) on COVID-19 outcomes [5,6,13,14,15]. Anti- CD20 monoclonal antibodies are commonly used in treatment of hematologic malignancy, but there is little data regarding the impact of these agents on COVID-19 course.

To address this dearth of data, we performed a retrospective analysis of the potential effects of commonly used immunomodulatory agents on cancer patients with COVID-19.

## Methods

### Patients

The study included all adult cancer patients followed at Memorial Sloan Kettering Cancer Center (MSKCC) who developed COVID-19 in the inpatient or outpatient setting between March 8, 2020 and June 2, 2020. SARS-CoV-2 status was determined using a nasopharyngeal swab to determine the presence of virus specific RNA (MSKCC FDA EUA-approved assay and Cephied®). Clinical outcomes were monitored until June 3, 2020. All patient data was obtained from the MSKCC electronic medical record. The MSKCC institutional review board approved the study and waived the requirement for informed consent.

### Data sources

Patient data was extracted from the MSKCC electronic medical record (EMR). Patient medications, demographics, laboratory measurements, and outcomes (i.e. use of supplemental oxygen, noninvasive or invasive mechanical ventilation, and death), were extracted from a standardized-input institutional database. Relevant comorbidities as well as thoracic and hematologic cancer status were extracted from International Classification of Disease (ICD)-9 and ICD-10 codes using a process cross-validated with manual curation [7].

### Statistical analysis

For all analyses we considered the number of patients who developed a primary composite endpoint of respiratory failure (use of nonrebreather, high-flow nasal oxygen, or mechanical ventilation) or death within 28 days of SARS-CoV-2 diagnosis.

### High-dose Corticosteroids

We evaluated the impact of corticosteroid administration (defined as the equivalent of at least 60mg cumulative of prednisone over COVID-19 course) on the composite outcome. In an attempt to partially control for selection bias in which more acutely ill patients received corticosteroids with therapeutic intent, we performed a series of multivariate Cox proportional hazards analyses with hematologic malignancy and corticosteroid use as variables. Corticosteroid administration was introduced as a time-dependent variable. We performed a “pre-supplemental oxygen” analysis from time of SARS-CoV-2 diagnosis to an endpoint of first use of >2L/min supplemental oxygen or death. We also performed a “post-supplemental oxygen” analysis from a start time of first use of >2L/min supplemental oxygen to an endpoint of respiratory failure or death. This model included only patients who had received >2L/min supplemental oxygen. The 2L/min cutoff was chosen to cohort patients with similar levels of COVID-19 severity at time of corticosteroid administration. We also performed a “post-critical” analysis from a start time of first use of nonrebreather, high-flow nasal oxygen, or mechanical ventilation to an endpoint of death. All patient events for these analyses were right-censored at 28 days following SARS-CoV-2 diagnosis or June 3, 2020, whichever came first. Hazard ratios (HR) and 95% confidence intervals (CI) were reported for these analyses.

### Anti-CD20 Therapy

We estimated the incidence in composite outcome in patients receiving anti-CD-20 therapy (rituximab, ofatumumab, or obinutuzumab within 90 days prior to SARS-CoV-2 diagnosis). Because patients with hematologic malignancy are more likely to receive these agents, we also estimated the rate of respiratory failure or death just among patients with hematologic malignancy.

### Immunotherapy

We estimated the incidence of the composite outcome in patients receiving immunotherapy (PD- 1, PDL-1, or CTLA-4 blockade within 90 days prior to SARS-CoV-2 diagnosis). This was additionally estimated separately among patients with a history of thoracic malignancy and among patients with metastatic disease and who had types of cancer that might make them eligible for immunotherapy, as performed by previous studies [6].

### Neutropenia

We evaluated the association of “pre-COVID-19 neutropenia” with the composite outcome. Pre-COVID-19 neutropenia was defined as any absolute neutrophil count, ANC < 1 K/uL between 7 and 60 days prior to SARS-CoV-2 diagnosis. We also performed analyses stratified based on whether or not a given patient received cytotoxic chemotherapy in the same time frame and whether or not the patient had a neutropenic measurement within 30 days of chemotherapy administration. We also considered whether or not a given patient had a diagnosis of hematologic malignancy. We separately examined incidences of respiratory failure or death in patients with “mild neutropenia,” i.e. at least one recorded ANC < 2 K/uL between 7 to 60 days prior to SARS-CoV-2 diagnosis but no recorded ANC < 1K/uL in that timeframe. We also examined incidences of respiratory failure or death in patients with “recovered neutropenia,” i.e. at least one ANC < 1K/uL in a given time window prior to SARS-CoV-2 diagnosis but not subsequently. For the “recovered neutropenia” analysis, two time windows were considered: 7 to 60 days and, separately, 60 to 180 days prior to SARS-CoV-2 diagnosis.

We compared interleukin-6 (IL-6), C-reactive protein (CRP), ferritin, and white blood cell (WBC) levels prior to and after high-dose corticosteroid administration using Wilcoxon signed-rank tests. Only values seven days prior to or after medication administration were considered. In the case of repeat laboratory measurements within the seven-day window the value closest to medication administration was used. We also compared those laboratory measurements within three days of SARS-CoV-2 diagnosis between patients who did and did not receive prior immunotherapy using Mann-Whitney U tests.

All analyses were performed using Python 3.7.4 and the “lifelines” package version 0.22.8.

## Results

A total of 820 cancer patients were diagnosed with SARS-CoV-2 during the study period. Patient characteristics are reported in Table 1. Collinearity between these characteristics is explored in Figure S1. A total of 761 patients (92.8%) were followed for the maximum possible time of 28 days or died. A total of 382 patients (46.5%) were admitted, 154 (18.8%) reached the primary composite endpoint, and 97 (11.8%) died.

**Table 1.**
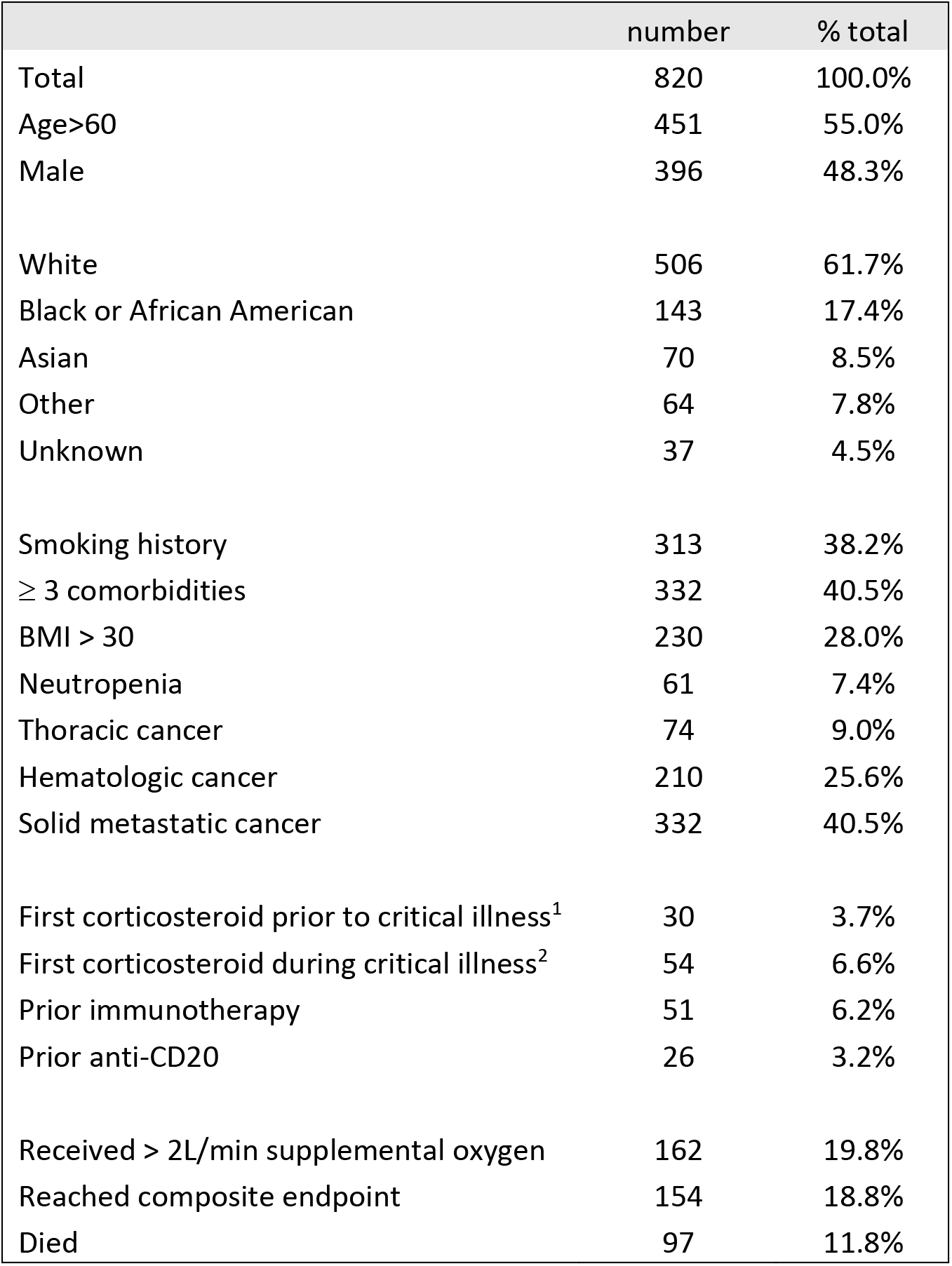
Patient characteristics (not mutually exclusive). ^1^High-flow oxygen, ventilation, or death. ^2^During high-flow oxygen or ventilation and prior to death.

|  | number | % total |
| --- | --- | --- |
| Total | 820 | 100.0% |
| Age>60 | 451 | 55.0% |
| Male | 396 | 48.3% |
| White | 506 | 61.7% |
| Black or African American | 143 | 17.4% |
| Asian | 70 | 8.5% |
| Other | 64 | 7.8% |
| Unknown | 37 | 4.5% |
| Smoking history | 313 | 38.2% |
| ≥ 3 comorbidities | 332 | 40.5% |
| BMI > 30 | 230 | 28.0% |
| Neutropenia | 61 | 7.4% |
| Thoracic cancer | 74 | 9.0% |
| Hematologic cancer | 210 | 25.6% |
| Solid metastatic cancer | 332 | 40.5% |
| First corticosteroid prior to critical illness<sup>1</sup> | 30 | 3.7% |
| First corticosteroid during critical illness<sup>2</sup> | 54 | 6.6% |
| Prior immunotherapy | 51 | 6.2% |
| Prior anti-CD20 | 26 | 3.2% |
| Received > 2L/min supplemental oxygen | 162 | 19.8% |
| Reached composite endpoint | 154 | 18.8% |
| Died | 97 | 11.8% |

Patients with a history hematologic malignancy were more likely to receive corticosteroids (Figure S1). Among patients with hematologic malignancy, 15 patients received corticosteroids, seven (48%) of whom developed respiratory failure or died. By contrast 52 of 195 (27%) patients with hematologic malignancy not receiving steroids reached that endpoint. These medications were also administered with higher levels of supplemental oxygen relative to patients who did not develop respiratory failure or died (Figure S2). When patients were stratified by level of respiratory support, corticosteroid use was associated with worse outcomes in the pre-2L oxygen cohort (HR 2.3, 95% CI 1.1-4.9), a trend not observed in the post-2L oxygen (HR 0.9, 95% CI 0.4-1.9) and post-critical cohorts (HR 0.8, 95% CI 0.5-1.4), though these additional analyses were limited by the sample size (Table S1). Indications for corticosteroid administration in the pre-hypoxic setting are reported in Table S2; there was no clear link between steroid indication and outcomes. Five of 17 (29%) patients who received high-dose corticosteroids in the pre-hypoxic setting had a prior standing prescription for corticosteroids at the time of COVID-19 diagnosis; of those five patients, two ultimately utilized >2L/min oxygen. IL-6 and CRP levels were lower, while ferritin levels were higher, after corticosteroid administration in both the pre and post-critical illness settings (Figure S3).

Nine of 26 patients (35%) treated with prior anti-CD20 agents had respiratory failure or died; although this was higher than the rate of respiratory failure or death in the cohort as a whole, the rates based on prior anti-CD20 agents were similar in the subset of patients with hematologic cancer (Table 2). The rates of respiratory failure or death based on prior immunotherapy were estimated in both the subset of patients with metastatic thoracic cancer and the subset of other metastatic solid cancers that were eligible for immunotherapy (Table 2). Nine of 25 patients (35%) with thoracic metastatic cancer who received immunotherapy developed respiratory failure or died, compared with 10 of 31 (32%) patients with thoracic metastatic cancer who did not receive immunotherapy. Immunotherapy administration was not associated with elevated IL-6 and CRP at time of SARS-CoV-2 diagnosis (Figure S3).

**Table 2.** Rate respiratory failure or death among patients treated and not treated with anti-CD20 agents and immunotherapy (IO) and with or without neutropenia pre-COVID-19. ^1^Limited to patients with solid metastatic cancer deemed eligible for immunotherapy by cancer type (N=179), specifically triple negative breast cancer, colorectal cancer, renal cancer, bladder cancer, melanoma, cervical or uterine cancer, esophageal or stomach cancer, or hepatocellular carcinoma. ^2^Mild neutropenia: at least one measured ANC < 2K/uL and all measured ANC > 1K/uL within 7 to 60 days prior to SARS-CoV-2 diagnosis ^3^Recovered neutropenia: at least one measured ANC<1K/uL during the stated time window and subsequently at least one ANC>1K/uL and no measurements of ANC<1K/uL after SARS-CoV-2 diagnosis.

|  | # resp. failure or death / total in category |  |
| --- | --- | --- |
|  | Received anti-CD20 | No anti-CD20 |
| Total | 9 / 26 (35%) | 145 / 794 (18%) |
| Hematologic | 7 / 23 (30%) | 52 / 187 (28%) |
| Solid | 2 / 3 (66%) | 93 / 607 (15%) |
|  | Received IO | No IO |
| Total | 14 / 51 (27%) | 140 / 769 (18%) |
| Thoracic (all) | 9 / 26 (35%) | 12 / 48 (25%) |
| Other solid (all) | 4 / 23 (17%) | 79 / 511 (15%) |
| Thoracic (metastatic) | 9 / 25 (36%) | 10 / 31 (32%) |
| Selected solid (metastatic)<sup>1</sup> | 3 / 18 (17%) | 18 / 101 (18%) |
|  | Neutropenia | No neutropenia |
| Total | 24 / 61 (39%) | 130 / 759 (17%) |
| On cytotoxic chemotherapy | 14 / 38 (37%) | 33 / 180 (18%) |
| No chemotherapy | 10 / 23 (43%) | 97 / 579 (17%) |
| Hematologic | 15 / 37 (41%) | 44 / 173 (25%) |
| Solid | 9 / 24 (38%) | 86 / 586 (15%) |
| Mild neutropenia<sup>2</sup> | 18 / 107 (17%) | 112 / 652 (17%) |
| 7-60 days pre-COVID, recovered<sup>3</sup> | 15 / 35 (43%) | 139 / 785 (18%) |
| 60-180 days pre-COVID, recovered | 4 / 24 (17%) | 150 / 796 (19%) |
|  | Received corticosteroid | No corticosteroid |
| Total | 10 / 30 (33%) | 144 / 790 (18%) |
| Hematologic | 7 / 15 (47%) | 52 / 195 (27%) |
| Solid | 3 / 15 (20%) | 92 / 595 (15%) |

Cytotoxic chemotherapy administration seven to 60 days prior to SARS-CoV-2 diagnosis was not associated with worse outcomes; however, patients who developed neutropenia following cytotoxic chemotherapy had worse outcomes. Of 38 patients with chemotherapy-associated pre-COVID-19 neutropenia, 14 (37%) developed respiratory failure or died. Patients with neutropenia unrelated to chemotherapy also had worse outcomes, regardless of solid or hematologic cancer type (Table 2). Of note, patients with neutropenia seven to 60 days prior to SARS-CoV-2 diagnosis but had “recovered” from neutropenia also had worse outcomes. Patients who had recovered from neutropenia 60 to 180 days prior to SARS-CoV-2 diagnosis did not have worse outcomes. Patients with mild neutropenia only (ANC < 2K/uL) did not have worse outcomes (Table 2).

Of note, four patients had pre-COVID-19 neutropenia and were treated with immunotherapy; of these four patients, three developed respiratory failure or died.

## Discussion

We present a single-center, retrospective analysis of cancer patients with COVID-19 treated with various immunomodulatory agents. Our study demonstrates that in assessing whether certain oncologic medications are associated with worse COVID-19 outcomes, considering cancer type, degree of effect (i.e. neutropenia or other bone marrow suppression) and other patient-specific factors is crucial.

Studies in non-cancer patients have suggested that corticosteroids may have a role in early COVID-19 illness [16]. In the cohort of patients treated with corticosteroids prior to use of >2L/min supplemental oxygen, we found a trend toward worse outcomes, although this may reflect selection bias for patients suspected at risk for decompensation receiving these medications. There is also promising evidence that corticosteroids may reduce mortality in critically ill patients with COVID-19-related respiratory failure [12,17]. We did not observe a decrease in adverse events in critically ill patients [12], although our cohort sizes for these subsets were limited. Further study on the impact of corticosteroids on critically ill cancer patients is needed. The clinical significance of lower IL-6 and CRP levels and higher ferritin levels following corticosteroid use in cancer patients also requires further inquiry.

We found that although cytotoxic chemotherapy itself was not a risk for worse outcomes, pre-COVID-19 neutropenia was associated with worse COVID-19. A minority of patients in our cohort receiving cytotoxic chemotherapy developed neutropenia; differences in the proportion of patients developing neutropenia while receiving chemotherapy may account for why previous studies have found both absence [4,5] and presence [8] of harm with these agents. Previous trends toward worse COVID-19 outcomes with thrombocytopenia at baseline and during infection have also been observed [7]. At the same time, abnormally high ANC has also been observed during in certain patients with worse COVID-19 [7]. It is plausible that bone marrow suppression or other immunosuppressive features, rather than neutropenia per se, are responsible for the worse outcomes seen in the neutropenic cohort of our study. It is noteworthy that even patients who may be recovering from relatively recent neutropenic events were found to have worse COVID- 19 outcomes. Although it is reassuring that more distant neutropenia and milder neutropenia were not associated with adverse outcomes, the relationship between recovery from immunosuppression and COVID-19 warrants further exploration.

The observed rates of respiratory failure or death were 36% and 32% for metastatic lung cancer patients who did and did not receive immunotherapy, respectively. We utilized a definition of metastatic disease that includes only solid tumors with distal or widespread metastases which may in part account for the variable findings compared to previous report in a similar population [6]. Immunotherapy use, thoracic cancer, smoking history, and metastatic solid cancer are highly associated (Figure S1), making it difficult to truly understand which are truly responsible for worse COVID-19 outcomes. Evaluation of the specific risks of immunotherapy use in patients with COVID likely requires larger analyses in cancer-specific cohorts.

It is also notable that of patients with pre-COVID-19 neutropenia treated with immunotherapy, three of four developed respiratory failure or died. All of these patients had solid metastatic disease and were treated with either prior or concomitant cytotoxic chemotherapy. Although recent chemotherapy is currently not believed to be a risk factor for worse COVID-19 outcomes, it has been observed that patients receiving combination chemotherapy and immunotherapy may have higher rates of severe respiratory COVID-19 [7]. The potential interaction between antineoplastic agents as well as cancer severity and COVID-19 warrants further study.

Despite efforts to control for potential confounders, this study has several limitations inherent to retrospective research. Because of the modest number of patients treated with most of the medications in question, more complex multivariate models were not performed. Certain immunomodulatory agents could not be included due to sparse representation. For example, 12 patients in our cohort received intravenous immunoglobulin; five of these patients developed respiratory failure or died. Inferring the harm or benefit of medications such as corticosteroids in a retrospective study is inherently biased. It is plausible that sicker patients were more likely to receive corticosteroids, and that this was not captured by our study variables. Many risk factors for COVID-19 are unknown, and COVID-19 treatment patterns are complex. It is certain that both patient baseline and COVID-19 related confounders exist in our dataset. Randomized, prospective cancer-specific clinical trials are needed to evaluate the safety and effectiveness of these agents in treating cancer patients with COVID-19.

## Data Availability

Data available upon request

## Disclosures

JJ has a patent licensed by MDSeq Inc. AJS holds shares in Aprea Therapeutics and Editas Medicine and his spouse works for and receives salary support from BluePrint Research Group and in the course of her position consults for multiple pharmaceutical companies. SAV has received honoraria from Agios Pharmaceuticals and Rheos Pharmaceuticals, is an advisor for Immunai and has consulted for ADC Therapeutics. SP has received research funding from the Marcus Foundation and from bioMerieux. NK has received research funding from Amgen and has served on an advisory board for Astra Zeneca. MS has served as a paid consultant for McKinsey & Company, Angiocrine Bioscience, Inc., and Omeros Corporation. He received research funding from Angiocrine Bioscience, Inc. He served on an ad hoc advisory board for Kite – A Gilead Company. SH has received research support from ADC Therapeutics, Aileron, Celgene, Daiichi Sankyo, Forty Seven, Inc., Kyowa Hakko Kirin, Millennium/Takeda, Seattle Genetics, Verastem, Portola Pharmaceuticals, and Trillium Therapeutics and has served as a consultant for ADC Therapeutics, C4 Therapeutics, Janssen, Kura Oncology, Kyowa Hakko Kirin, Myeloid Therapeutics, Seattle Genetics, Takeda, and Trillium Therapeutics.

## Funding

This research was funded in part through the NIH/NCI Cancer Center Support Grant P30 CA008748 and by the Memorial Sloan Kettering Cancer Center K12 Paul Calabresi Career Development Award for Clinical Oncology (to A.D.).

---

**Table S1.** HR and 95% CI in multivariate analysis for 1. Analysis of time from SARS-CoV-2 diagnosis to use of >2L/min supplemental oxygen or death and 2. Analysis from time of >2L/min supplemental oxygen use to need for high-flow oxygen, intubation, or death. 3. Analysis from time of high-flow oxygen or intubation to death.

|  | Pre-supplemental oxygen |  |  | Post-supplemental oxygen |  |  | Post-critical |  |  |
| --- | --- | --- | --- | --- | --- | --- | --- | --- | --- |
|  | HR | - 95%CI | + 95%CI | HR | - 95%CI | + 95%CI | HR | - 95%CI | + 95%CI |
| Hematologic cancer | 1.6 | 1.2 | 2.0 | 1.4 | 0.9 | 2.0 | 0.9 | 0.5 | 1.4 |
| Corticosteroid event | 2.3 | 1.1 | 4.9 | 0.9 | 0.4 | 1.9 | 0.8 | 0.5 | 1.4 |

**Table S2.** Indications for high-dose corticosteroid use in the pre-supplemental oxygen setting. “Event” signifies progression to utilize >2L/min supplemental oxygen.

| Steroid Indication | No event (N=10) | Event (N=7) | Total (N = 17) |
| --- | --- | --- | --- |
| COVID-19 treatment | 2 | 3 | 5 |
| Suspected adrenal insufficiency | 3 | 2 | 5 |
| Cancer treatment | 1 | 2 | 3 |
| Immunotherapy toxicity | 1 | 0 | 1 |
| Other | 3 | 0 | 3 |

**Figure S1.**
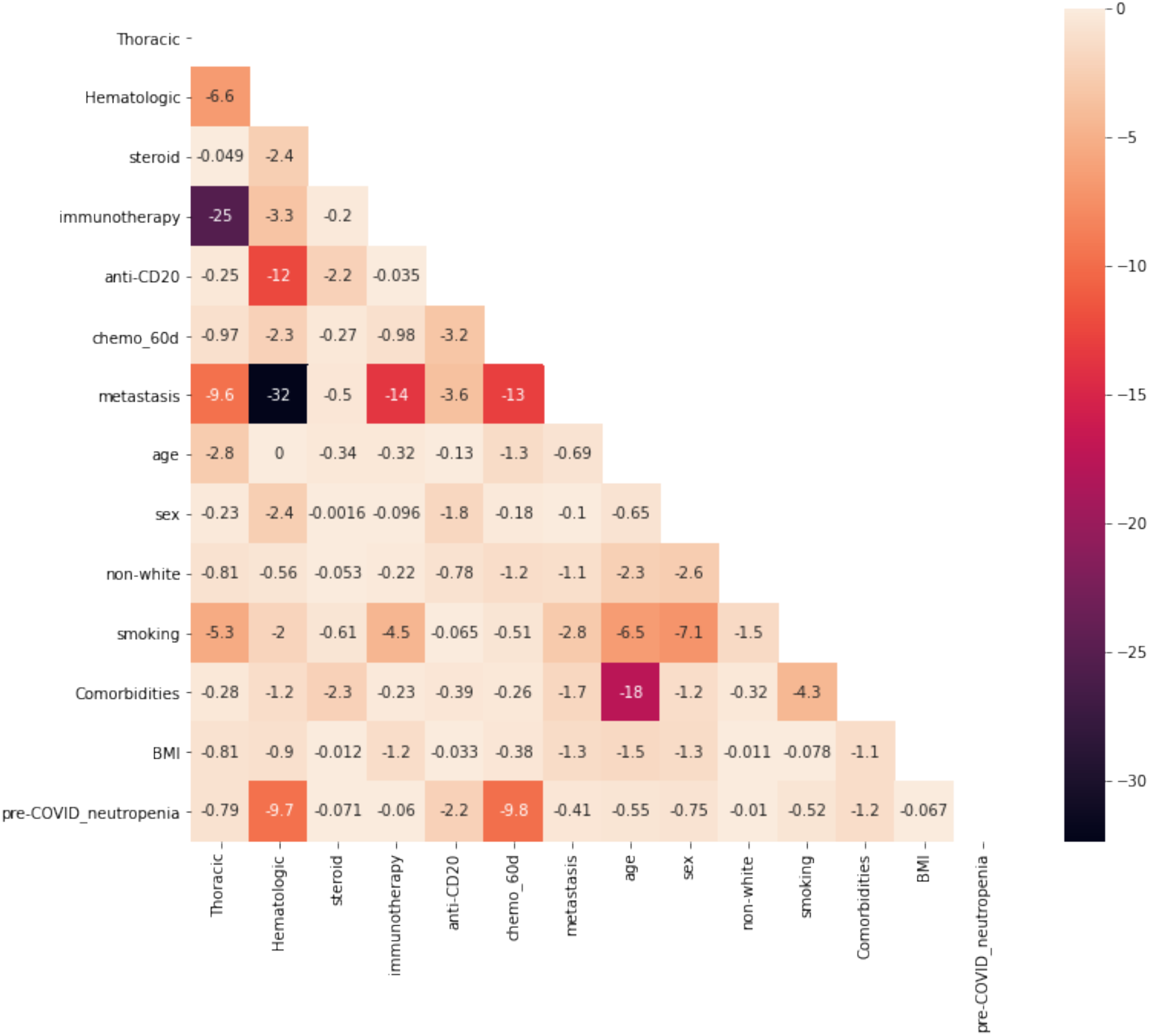
Pairwise Chi-square testing among variables. Values shown are log_10_(p-value). chemo_60d = cytotoxic chemotherapy within 60 days of SARS-CoV-2 diagnosis.

**Figure S2.**
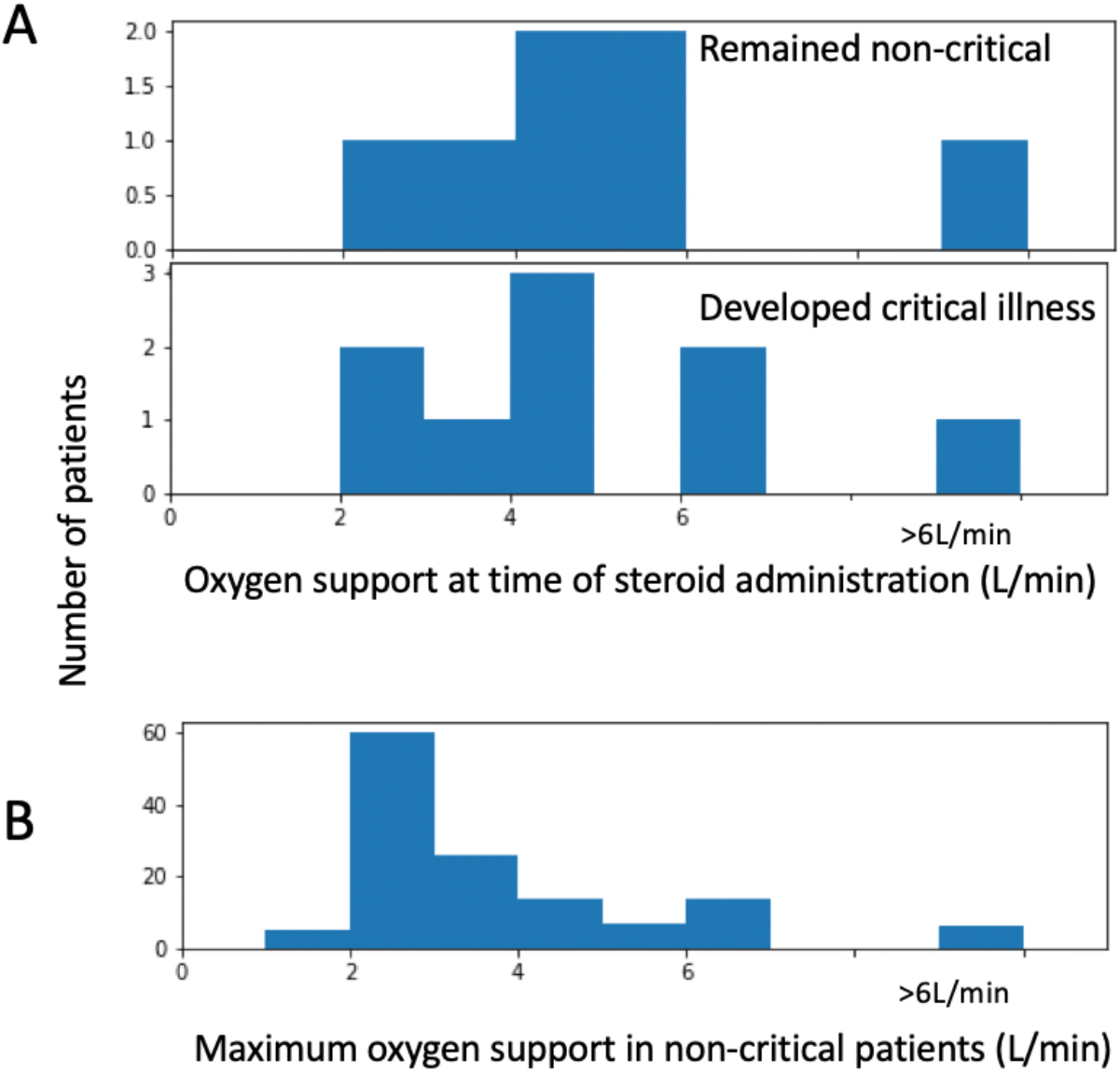
A. Histogram of amount of oxygen (L/min) a given patient was on when receiving highdose corticosteroids, separated into patients who developed a requirement for high-flow oxygen or mechanical ventilation or died (“Developed critical illness”) or did not (“Remained non-critical”). B. Histogram of maximum oxygen support in all patients who utilized supplemental oxygen but did not develop critical illness. Patients who did not require oxygen or who ultimately received high-flow oxygen or mechanical ventilation are excluded.

**Figure S3.**
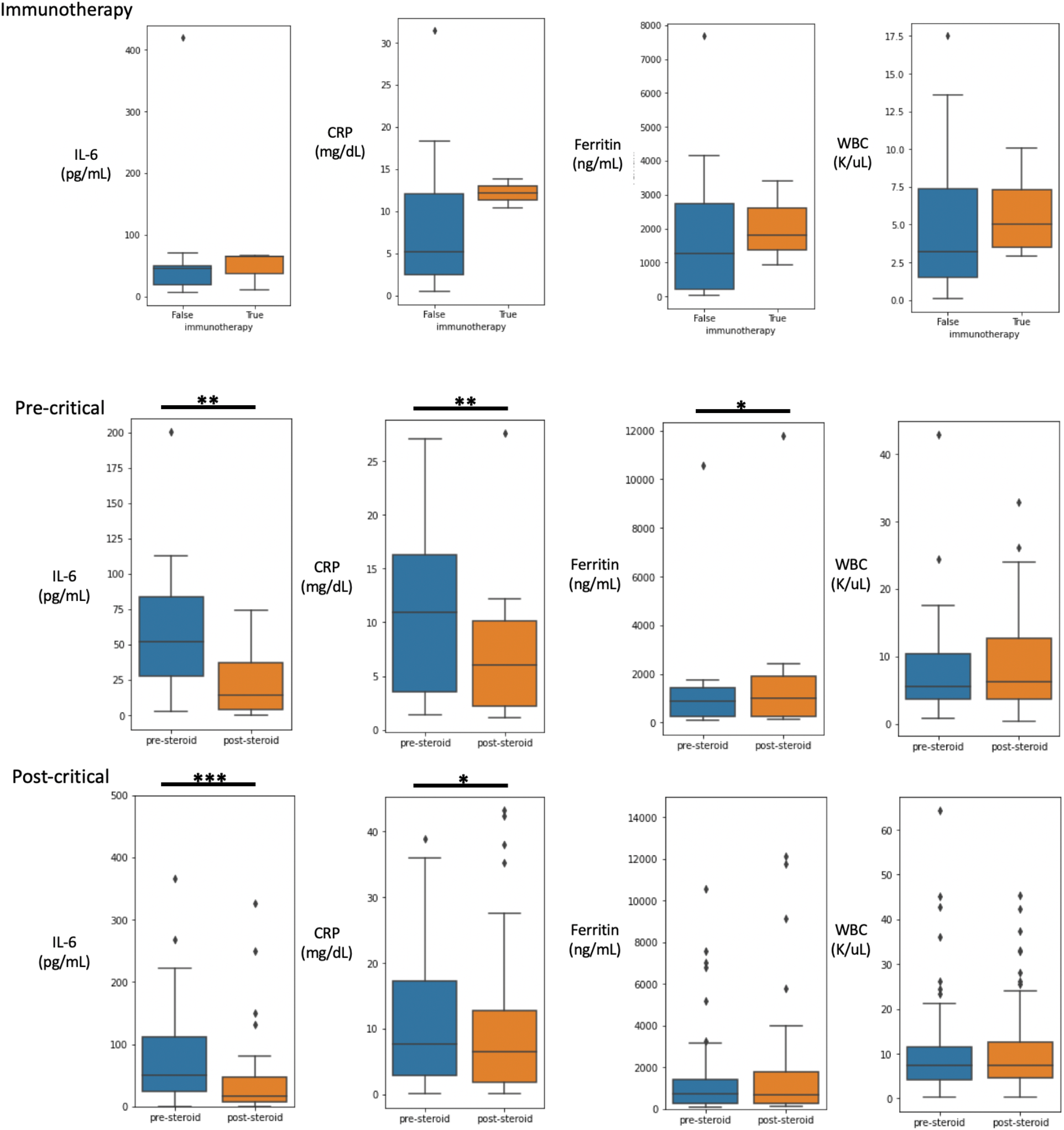
(Top) Boxplot showing quartiles and outliers of earliest laboratory values within 3 days of SARS-CoV-2 diagnosis for patients treated or not treated with immunotherapy 90 days prior. No comparisons were significant by Mann-Whitney U test with p-value cutoff of 0.05. Below, boxplots showing quartiles with outliers of paired laboratory values from prior to and after administration of high-dose corticosteroids (middle) prior to respiratory failure (nonrebreather, high-flow nasal oxygen or mechanical ventilation) and (bottom) after respiratory failure. In the post-critical IL-6 plot, outliers >500pg/mL are excluded. In the post-critical ferritin plot, outliers > 15,000ng/mL are excluded. *** p< 0.0001, ** p<0.001, * p<0.01 by Wilcoxon signed-rank test.

## Notes

### Author Declarations

This study was approved by the Memorial Sloan Kettering Institutional Review Board.

## References

[1] Wang, D. et al. Clinical characteristics of 138 hospitalized patients with 2019 novel coronavirus-infected pneumonia in Wuhan, China. JAMA 323, 1061-1069 (2020).

[2] Dai M, et al. Patients with cancer appear more vulnerable to SARS-COV-2: a multi-center study during the COVID-19 outbreak. Cancer Discov. CD-20–0422. https://doi.org/10.1158/2159-8290.CD-20-0422. (2020)

[3] Mehta, V, et al. Case Fatality Rate of Cancer Patients with COVID-19 in a New York Hospital System. Cancer Discovery. DOI: 10.1158/2159-8290.CD-20-0516 (2020)

[4] Kuderer, N. M. et al. Clinical impact of COVID-19 on patients with cancer (CCC19): a cohort study. The Lancet. https://doi.org/10.1016/S0140-6736(20)31187-9 (2020)

[5] Lee, L.Y.W. et al. COVID-19 mortality in patients with cancer on chemotherapy or other anticancer treatments: a prospective cohort study. The Lancet. https://doi.org/10.1016/S0140-6736(20)31173-9 (2020)

[6] Robilotti, E.V et al. Determinants of Severity in Cancer Patients with COVID-19 Illness. Nature Medicine. https://doi.org/10.1038/s41591-020-0979-0 (2020)

[7] Jee, J. et al. Chemotherapy and COVID-19 Outcomes in Patients with Cancer. Journal of Clinical Oncology. Forthcoming (2020)

[8] Yang, K. et al. Clinical characteristics, outcomes, and factors for mortality in patients with cancer and COVID-19 in Hubei, China: a multicenter retrospective cohort study. Lancet Oncol. Doi:10.1016/S1470-2045(20)30310-7 (2020)

[9] WHO. Clinical management of severe acute respiratory infection when novel coronavirus (nCoV) infection is suspected. Geneva, World Health Organization, https://www.who.int/internal-publications-detail/clinical-management-of-severe-acute-espiratory-infection-when-novelcoronavirus-(ncov)-infection-is-suspected (2020)

[10] IDSA. Infectious Disease Society of America Guidelines on the Treatment and Management of Patients with COVID-19. April 11. https://www.idsociety.org/practice-guideline/covid-19-guideline-treatment-and-management/ (2020)

[11] Rivera, D.R et al. Utilization of COVID-19 treatments and clinical outcomes among patients with cancer: A COVID-19 and Cancer Consortium (CCC19) cohort study. Cancer Discovery. Doi:10.1158/2159-8290.CD-20-0941 (2020)

[12] The RECOVERY Collaborate Group. Dexamethasone in Hospitalized Patients with Covid-19—Preliminary Report. NEJM. Doi: 10.1056/NEJMoa2021436 (2020)

[13] Abid, M.B et al. Coronavirus Disease 2019 (COVID-19) and Immune-Engaging Cancer Treatment. JAMA Oncol. Doi:10.1001/jamaoncol.2020.2367 (2020)

[14] Luo, J. et al. Impact of PD-1 blockade on severity of COVID-19 in patients with lung cancers. Cancer Discovery. DOI: 10.1158/2159-8290.CD-20-0596 (2020)

[15] Lara, O.D et al. COVID-19 Outcomes of Gynecologic Cancer Patients in New York City. Cancer. Forthcoming (2020)

[16] Fadel, R D, et al. COVID-19 Management Task Force, Early Short Course Corticosteroids in Hospitalized Patients with COVID-19, Clinical Infectious Diseases, doi.org/10.1093/cid/ciaa601 (2020)

[17] Villar, J. et al. Rationale for Prolonged Corticosteroid Treatment in the Acute Respiratory Distress Syndrome cause by Coronavirus Disease 2019. Crit Care Expl 2:e0111 (2020)

